# Predicting COVID-19 booster immunogenicity against future SARS-CoV-2 variants and the benefits of vaccine updates

**DOI:** 10.1101/2024.02.08.24302032

**Authors:** Deborah Cromer, Arnold Reynaldi, Ainslie Hie, Timothy E. Schlub, Jennifer A. Juno, Adam K. Wheatley, Stephen J. Kent, David S. Khoury, Miles P. Davenport

## Abstract

The ongoing evolution of the SARS-CoV-2 virus has led to a move to update vaccine antigens in 2022 and 2023. These updated antigens were chosen and approved based on in vitro neutralisation titres against recent SARS-CoV-2 variants. However, unavoidable delays in viral manufacture and distribution meant that the updated booster vaccine was no longer well matched to the circulating SARS-CoV-2 variant by the time of its deployment. Understanding whether the updating of booster vaccine antigens improves immune responses to subsequent SARS-CoV-2 circulating variants is a major priority in justifying future vaccine updates. Here we analyse all available data on the immunogenicity of variant containing SARS-CoV-2 vaccines and their ability to neutralise later circulating SARS-CoV-2 variants. We find that updated booster antigens give a 1.4-fold [95%CI 1.07-1.82] greater increase in neutralising antibody levels when compared with a historical vaccine immunogen. We then use this to predict the relative protection that can be expected from an updated vaccine even when the circulating variant has evolved away from the updated vaccine immunogen. These findings help inform the roll out of future booster vaccination programs.

## Introduction

The continual emergence of novel SARS-CoV-2 variants has driven vaccine manufacturers and regulatory bodies to update COVID-19 vaccine immunogens^29,30^. Ideally, the choice of which vaccine to deploy would be informed by vaccine efficacy trials against the relevant circulating SARS-CoV-2 strains. However, the decision of which immunogen to include in a vaccine occurs well before a vaccine’s eventual approval, manufacture, and deployment, and therefore randomised controlled trials are not possible in the time required to inform strain selection decisions^31^. As a result, the assessment and comparison of different updated vaccine immunogens has been made based on neutralising antibody titres after vaccination, most commonly against the vaccine immunogen^21,22,32,33^. Since continued evolution and neutralization escape can occur before an updated vaccine is widely administered, in general the circulating variant at the time of vaccine deployment no longer matches the immunogen that was included in the vaccine and against which the vaccine was assessed^34^.

The advantage of an updated to a vaccine immunogen therefore must be assessed in the context of an anticipated mismatch between the vaccine and the circulating strains. The relative benefits of updating a vaccine immunogen are thought to be dependent on the antigenic relationships between the historical, contemporaneous and the future circulating variants. That is, the infection and vaccination history of an individual are thought to contribute to their current cross-recognition profile of different variant strains^35^. Further, boosting with a variant antigen leads to increased recognition of the vaccine strain (and antigenically similar strains)^3^.

How effective this is at improving responses to a future variant is thought to be dependent on the similarity between the booster antigen and a future circulating variant (antigenic distance)^36^. However, this relationship to the future circulating variant is inherently unknowable at the time when the decision on which immunogen to include in a vaccine needs to be made. An alternative approach is to look to past experience to inform the likely future outcomes. That is, we can look to past examples and existing data to ask what (average) advantage updating the vaccine immunogen gave to recognition of a future variant?

In this work, we identified all available comparative studies of different vaccine immunogens and considered the reported in vitro neutralisation titres against a variant that primarily circulated during/after deployment of the vaccine (defined in Figure 2). We compared pairs of potential immunogens at a given time, one designated as ‘old’ and one as ‘updated’, and assessed their immunogenicity against a future variant (i.e. against a variant that circulated *chronologically after* the variant immunogen that was used in either the old or updated vaccine). This allowed to identify the degree to which, for a given variant wave, an ‘updated’ booster provided a better boost to the neutralising antibody titres of a future variant than an ‘old’ booster. We then used this data, and the established relationship between neutralising antibody response and protection from COVID-19, to predict the additional protection from COVID-19 disease that could be achieved by using an updated vaccine immunogen.

## Methods

### Data Acquisition

We searched the VIEW-hub ongoing systematic review database for papers reporting neutralising antibody titres against COVID-19 after vaccination with a COVID-19 variant-immunogen^37^ and indexed prior to 15 August 2023. This search identified 399 studies for screening. We also identified 6 presentations made to the FDA and one paper that was related to one of the papers identified through VIEW-hub, and screened those as well, leading to the screening of 406 papers or presentations.

We screened for studies that were annotated as either (i) including vaccines that contained a BA.1, BA.5, XBB or bivalent immunogen, or (ii) reporting neutralisation titres against the XBB variant. This second screening criteria ensured that we captured all later studies where non-ancestral vaccine immunogens may have been used but where the study may not have been correctly annotated by VIEW-hub. We removed 354 studies from further screening as they did not meet the criteria outlined above.

For studies to be included in our analysis, they must have reported neutralisation titres after boosting, with at least one variant containing immunogen. Studies were excluded if they (i) did not report data after boosting with a variant immunogen, (ii) reported data included in another publication, (iii) did not report neutralisation titres against a variant that occurred chronologically after the vaccine immunogen, (iv) did not specify which immunogen was included in the booster dose, or (v) did not include data from human subjects.

After screening, we identified a total of 23 of the remaining 52 publications that met our inclusion criteria (left side of Figure 1). We also included data from a further 5 studies (right side of Figure 1), that had previously been identified by us to include neutralisation data after boosting with variant modified vaccines in an earlier analysis^38^ and remained relevant for this analysis, leading to a total of 28 studies from which we could extract data relevant to this analysis (Figure 1). Full details of the studies identified are given in Supplementary Table 1.

**Figure 1.**
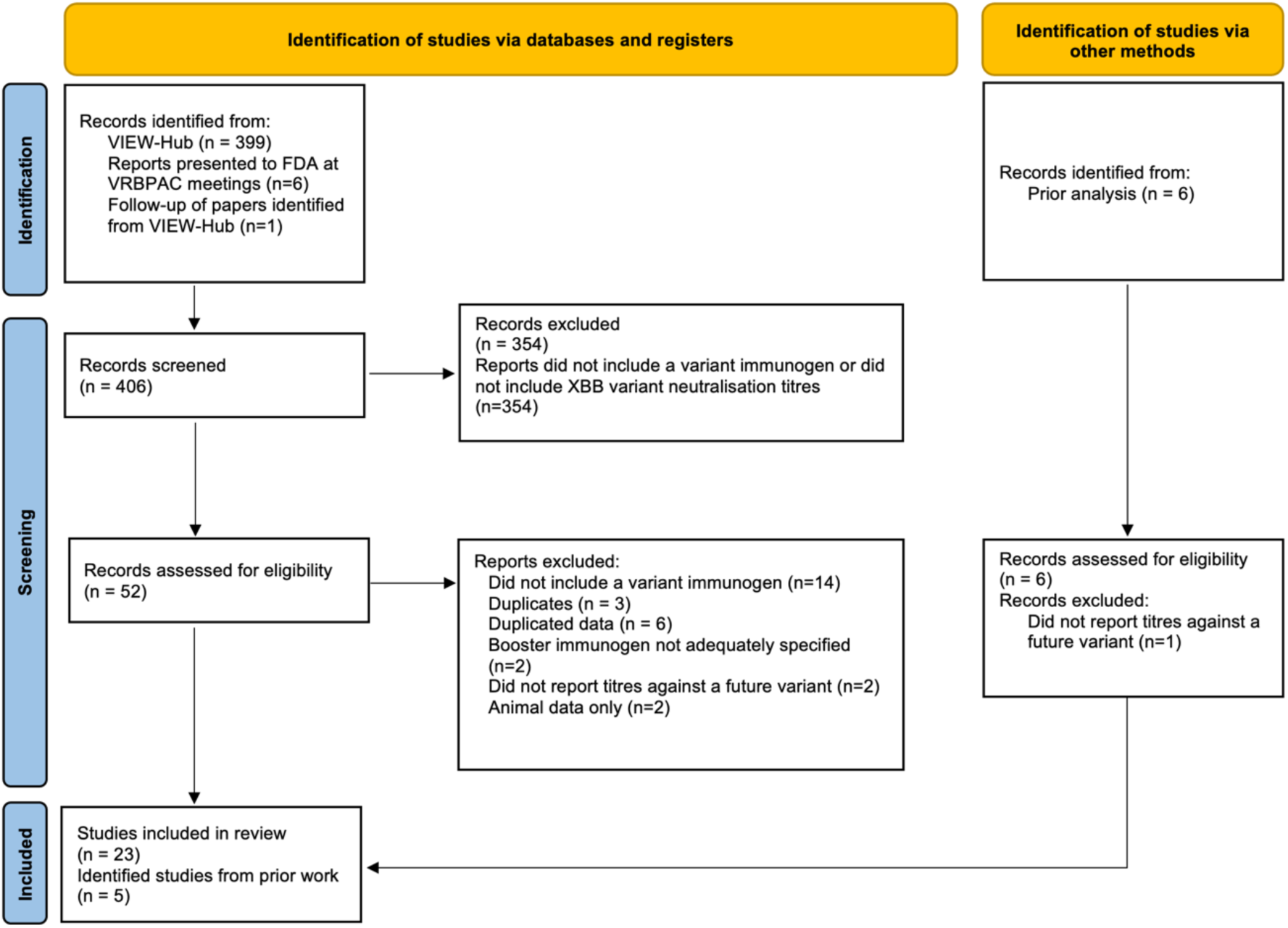
PRISMA Flow Chart showing studies selected for inclusion in the meta-analysis.

**Figure 2.**
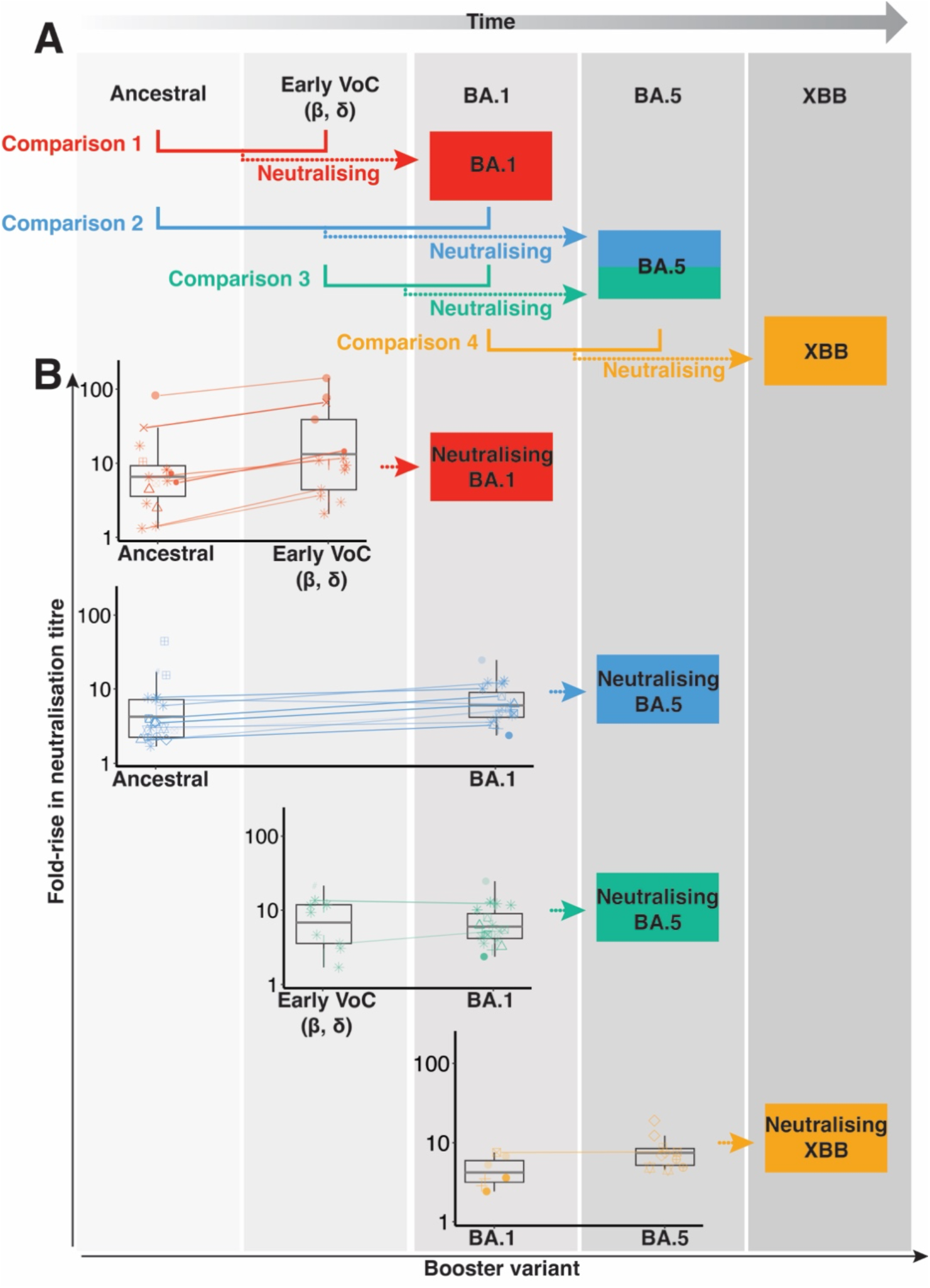
(A) Chronology of appearance of different SARS-CoV-2 variants and associated vaccine immunogens, and the comparisons made between different vaccine immunogens to induce neutralisation against future variants (coloured arrows). (B) Data extracted from identified papers that could contribute to each of the comparisons outlined in panel A. Lines connect paired cohorts from the same study.

### Data Extraction

We extracted neutralisation titres from the identified papers for cohorts:

1. That contained in vitro measurements against ancestral, BA.1, BA.5 and XBB variants.
2. That were unique (i.e. cohorts not already extracted from another paper).
3. Where studies reported outcomes disaggregated with respect to the status of prior infection (i.e. outcomes disaggregated into naïve and previously infected) prior to booster administration, we only extract this disaggregated data.
4. Where studies did not disaggregate data with respect to the status of prior infection, or where data was only disaggregated for either the naïve or previously infected portion of the cohort and was also presented for a mixed cohort. In this case, we extracted the neutralisation titres for the mixed cohort.
5. For which the most recent “boost” was by vaccination (not infection).
6. With subjects who had been given at least 2 prior doses of a vaccine.

We extracted neutralisation titres for the Ancestral, BA.1, BA.5 and XBB variants. Where a bivalent vaccine contained multiple immunogens (e.g. Ancestral + BA.5 or early VoC + BA.1) we classified the vaccine immunogen to be that of the latest chronological variant.

### Mixed Effects Model

We constructed a mixed effects model to determine if the neutralising antibody response following boosting (either the fold rise in neutralising antibodies or the absolute neutralising antibody titres following boosting) was dependant on the immunogen contained within the booster.

The model included a fixed effect for the booster immunogen (old or updated) and accounted for the study from which the data came by incorporating a random effect for the intercept with a grouping structure on the study from which the data came. It also accounted for the number of subjects within each cohort by weighing the contribution of each data point so that more weight was given to larger cohorts.

We also constructed an extended model that included the effects outlined above and additionally included a fixed effect for the exposure history of a cohort (average number of prior exposures within a cohort) and an additional random effect on the intercept with a grouping structure on the pairing of the data within a study (so that cohorts from the same study with the same exposure history, but with different booster immunogens, could be paired together within a study). The extended model also accounted for the comparison that was being made (i.e. comparison number 1, 2, 3 or 4 from Figure 2) as a grouping structure on the random effect for both the intercept and the booster immunogen. Full details of the model are provided in the supplementary materials.

## Results

### Identification of data from relevant studies

We identified 28 studies^1–28^ in which neutralising antibody titres were measured against future variants following variant-containing booster vaccination (Supplementary Table 1). From these we identified studies that considered in vitro neutralisation titres against BA.1, BA.5 and XBB variants, following booster vaccination with vaccines containing Ancestral, Beta, Delta, BA.1, BA.5 and XBB immunogens. Of these, 18 studies reported any pre-boost neutralisation titres (though in four of these studies pre-boost titres were not reported for all immunogen/variant combinations) and 10 studies only reported post-boost titres. Two studies analysed responses to a single relevant immunogen-containing booster, 20 compared neutralising antibodies after two different immunogen-containing boosters, and 6 compared responses after 3 or more immunogen-containing boosters.

### Immunogen Comparisons

Our aim was to consider whether boosting with an updated immunogen confers higher neutralising antibody titres to a future (at the time unknown) variant than boosting with an older immunogen. We identified four such comparisons that could be made from the available data, and these are shown by the coloured lines in Figure 2A.

Of the 28 studies we identified, only two studies^1,3^ contained data that could be used to contribute to all four comparisons, however no study contained data for *both* the old and updated immunogens across all four comparisons. In addition, 1 study could not be used in our analysis as it did not include titres after one of the relevant immunogens^9^, leaving 27 studies that could contribute to one or more of the comparisons identified in Figure 2A. Table 1 shows the number of studies we identified that could contribute to each comparison, and also highlights how many of these included pre-boost neutralisation titres, which could be used to calculate the fold-change in neutralisation titres after boosting. These fold-changes are depicted in Figure 2B.

**Table 1.** Number of studies reporting neutralisation titres against a future variant for each of the four possible comparisons. Data are split by those with data for either the old or updated immunogen and those with data for both the old and updated immunogens. Numbers in brackets give the number of studies that also included pre-boost neutralisation titres, and could therefore be included when modelling the fold-rise in neutralisation titres after boosting.

| **Comparison** | **<u>Comp 1</u>**<br>**Ancestral vs**<br>**Early VoC** | **<u>Comp 2</u>**<br>**Ancestral vs**<br>**BA.1** | **<u>Comp 3</u>**<br>**Early VoC vs**<br>**BA.1** | **<u>Comp 4</u>**<br>**BA.1 vs BA.5** |
| --- | --- | --- | --- | --- |
| **Neutralisation titres**<br>**against:** | **BA.1** | **BA.5** | **BA.5** | **XBB** |
| Either old or updated<br>immunogens | 15 (7) | 24 (13) | 11 (8) | 15 (7) |
| Both old and updated<br>immunogens | 5 (4) | 5 (4) | 1 (1) | 3 (1) |
| Benefit of update to<br>fold rise in<br>neutralisation titres<br>[95% CI] | 1.52<br>[1.27-1.81] | 1.68<br>[1.45-1.94] | 1.11<br>[0.88-1.41] | 1.19<br>[0.84-1.68] |

### Updated Immunogens Provide a Greater Boost to a Future Variant

We first investigated the relative benefit of an updated booster versus an older booster on neutralisation titres against the future variant (across the comparisons defined in Figure 2A). We constructed a mixed effects linear regression model that accounted for between study heterogeneity and the differences in the sizes of the cohorts and found that updated antigens consistently predicted better increases in neutralisation against the relevant variant.

The advantage of an updated booster ranged from a 1.11 to 1.68-fold increase in the fold-rise in neutralising antibody titres (Table 1). Considering all the data identified from studies that could contribute to the comparisons (56 cohorts from 16 studies), we found that the neutralisation titres against the specified future variant were, on average, 1.52-fold [95% CI 1.40-1.64] greater after boosting with the updated immunogen compared to the older immunogen.

The calculation above is subject to potential confounders. In particular, the comparisons being made (number 1, 2, 3 or 4 from Figure 2A) occurred in the context of different immune histories (eg: different numbers of prior vaccinations and / or infections, Supplementary Figure 1).

Therefore, we added these factors as co-variates in our model. We found that, consistent with previous work^39^, the overall level of boosting (of both old and updated vaccines) declined with an increasing number of prior exposures (by 2.5-fold per additional exposure [95%CI 2.1-3.1], Table 2). However, even after accounting for the number of prior exposures, boosting with an updated immunogen was estimated to give a 1.40-fold better boost than boosting with an older immunogen ([95%CI = 1.07-1.82], p=0.01). That is, on average, neutralisation titres to the future variant were boosted 40% higher after an updated immunogen boost, than after an older immunogen boost.

**Table 2.** Parameter values for the mixed effects model described in Equation S2, that predicts the fold-rise in neutralisation titres following boosting and is fit to the data shown in Supplementary Figure 1A.

| Dependant Variable | Parameter | Value | Std err | P-value |
| --- | --- | --- | --- | --- |
| Fold rise in neutralisation titres after boosting | Intercept | 1.245 | 0.089 | <0.001 |
|  | Immunogen (updated vs older) | 0.145 | 0.058 | 0.013 |
|  | Exposure number | -0.404 | 0.043 | <0.001 |

To assess the robustness of these observations we performed a range of sensitivity analyses. To confirm that cohorts with mixed prior infection status were not impacting our results, we repeated our analysis considering only uninfected cohorts or cohorts with homogeneous prior infection status (Supplementary Table 2 and Supplementary Table 3 respectively). We found that in both cases an updated immunogen also provided a greater boost to neutralisation titres (1.54 [95%CI 1.11-2.13] and 1.58 [95%CI 1.19-2.09] respectively).

Another potential confounder is that the early VoC immunogens (e.g. Beta and Delta containing vaccines) were never actually deployed in the population against COVID-19, unlike the other boosters immunogens we consider. To confirm that the comparisons incorporating early VoC vaccine immunogens were not biasing our results, we repeated the analysis excluding any comparison that involved an early VoC immunogen (ie: considering only comparisons 2 and 4). We found an almost identical benefit from the use of updated booster immunogen when excluding any comparisons that involved the early VoC immunogens (1.41-fold [95%CI 1.08-1.83] benefit of the updated immunogen, Supplementary Table 4).

Decisions on which immunogen to include in a vaccine are generally made on the basis of fold-rises in neutralisation titre after boosting (as described above). However, under 60% (16/27) of the studies we identified reported both pre- and post-boost titres necessary to calculate fold-rise in titre for the immunogen / variant combinations required). Therefore, to confirm that the actual neutralisation titres were improved by updating the vaccine immunogen (and not just the fold increase in neutralisation titres), we repeated the above analysis on the absolute neutralisation titres to a future variant reported after boosting. We found that the absolute neutralisation titres after boosting with an updated immunogen were estimated to be 1.52 fold [95%CI 1.30-1.77] higher than after boosting with an older immunogen (Supplementary Table 5).

Together, this demonstrates that booster vaccines containing chronologically more recent variant immunogens, have to date provided better neutralisation responses to the relevant future variant wave that circulated after their deployment.

### Optimal frequency for updating booster immunogens

The analysis above considers an updated booster immunogen against a recent alternative booster regimen. To date the booster immunogens have been updated annually^29,30^. However, it is possible to ask whether less frequent updating would achieve similar results. For example, in late 2022/early 2023, the XBB family variants were the dominant circulating strains, and there were three potential booster immunogens available for use; the original ancestral booster (dominant until mid-2021), an ‘older’ BA.1 booster (dominant late 2021 / early 2022), and an ‘updated’ BA.5 booster (dominant mid-2022). Using the available data for all three of these booster immunogens, we can ask whether repeated immunogen updates provided continued improvement to the boost in neutralisation titres.

When comparing in vitro neutralisation titres against XBB family variants we find that the update from the ancestral booster to a BA.1 booster improved the obtained boost in neutralisation titres, and the a further update from the BA.1 to a BA.5 vaccine immunogen gave an even better boost (Figure 3). From this, we can see that both the BA.1 and BA.5 variant immunogens gave a superior boost to XBB neutralisation titres than did the ancestral immunogen (Figure 3, in line with our previous work^38^) and in addition, that the updated BA.5 immunogen gave a better boost than the older BA.1 immunogen.

**Figure 3.**
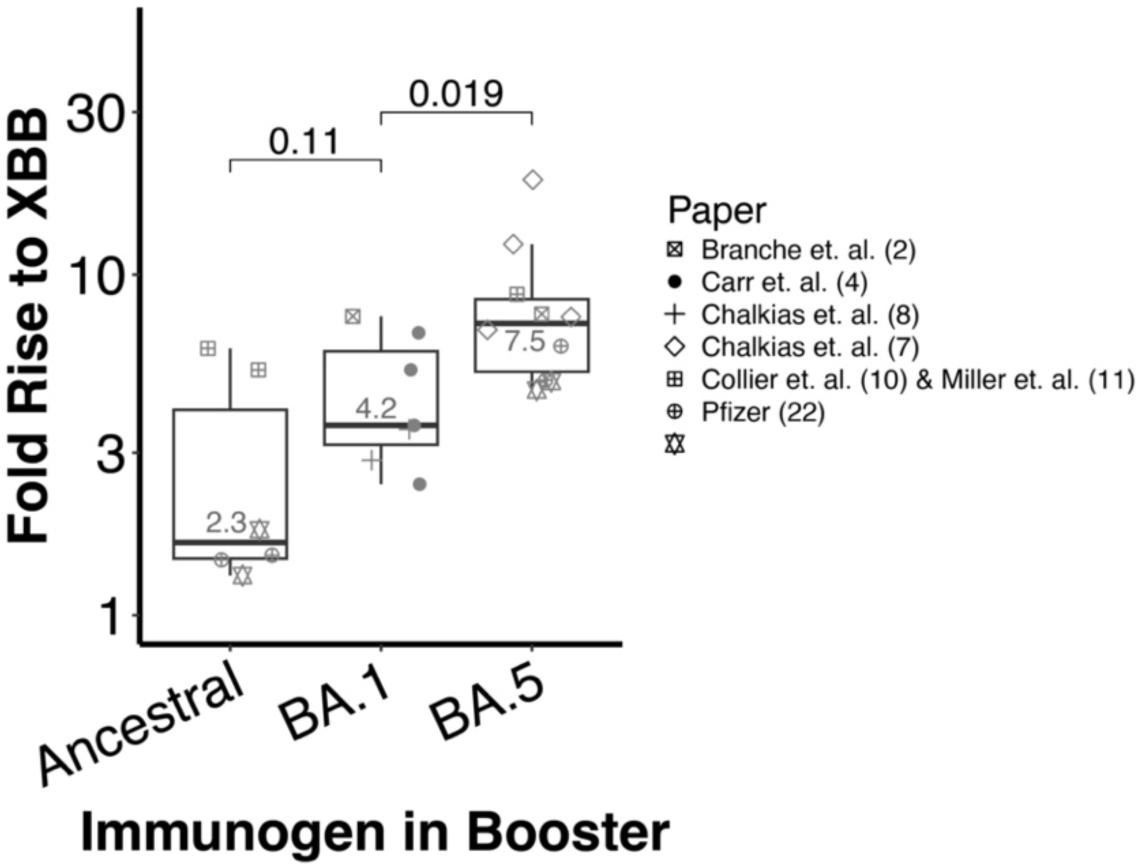
Fold rise in neutralisation titres to the XBB family variants after boosting with vaccines containing different booster immunogens. Small numbers show the geometric mean of the fold rises for each immunogen. Comparisons across the top show p-values from unpaired t-tests.

Together these observations suggest that if we were to stop updating booster antigens, then the current boosters would give a lower and lower boost to successive future variants. This suggests continued benefit for annual updating of the booster immunogen.

### Predicting the clinical benefit of updating the vaccine immunogen

Updating the immunogen in a booster vaccine appears to (on average) improve neutralisation titres by 40% to a future variant. However, it is not clear what this means for protection from disease. We have previously established a quantitative relationship between neutralizing antibody titres and protection from symptomatic and severe COVID-19^40,41^. We can use this relationship to estimate the additional odds of protection against symptomatic or severe infection that would result from boosting with an updated immunogen, compared to an old booster immunogen.

If an updated booster provides on average a 40% boost in titre compared to an older immunogen, then the odds ratio of protection from severe COVID-19 in a homogeneous population is 1.57 [95% CI 1.38-1.88]; i.e. the odds of protection from severe disease are increased by 57% compared to boosting with the old antigen. We can also consider how this might play out if we never updated immunogens. For example, if the benefit of updating is constant over different generations of booster antigen, as appears to be the case from Figure 3 (ie: a 40% increment for each update), then if an older booster vaccine is two immunogens behind (rather than only one), the odds ratio for severe disease in a population receiving the current booster, compared to one receiving a booster that is two immunogens behind, is predicted to be 1.57^2=2.46 [95% CI = 1.90-3.53].

How this translates into overall protection in a population is dependent on the background level of immunity and the protection that an old booster would provide^38^. We must therefore consider the predicted advantage of deploying an updated booster over and above the protection that would be achieved using an older booster vaccine. If an older booster vaccine gave rise to 60% protection in the population against symptomatic disease and 91% protection against severe disease, then the updated booster would be predicted to increase protection to 68% against symptomatic and 94% against severe disease. If a clinical trial were to be run, we would expect to observe a relative efficacy of 19% and 30% for the updated immunogen compared to the historical immunogen against symptomatic and severe disease respectively.

More generally, if we assume that an older booster offers between 30% and 80% protection against symptomatic disease, and an updated boosted immunogen gives a 1.4-fold improvement in neutralisation titres over this older vaccine, we would expect to observe an 11-25% effectiveness of the updated booster vaccine against symptomatic disease and a 23-33% effectiveness against severe disease compared to the older vaccine booster.

## Discussion

The rapid evolution of SARS-CoV-2 variants over time means that decisions on updating of booster immunogens must always be made with imperfect information. Here, we have assembled the (limited) data currently available to inform this decision. We find that the available evidence on previous booster comparisons suggests that updating of vaccine immunogens should provide an average of around a 40% greater boost to neutralisation titres against subsequent season’s SARS-CoV-2 variant, compared to a vaccine containing an immunogen with an older spike variant.

Interestingly, these results suggest that even if ‘updated’ vaccines are not rolled out quickly enough to immunise a population against the currently circulating variant, there remains an immunological benefit to boosting with a new, and possibly divergent antigen. Understanding the mechanisms by which booster immunisation can drive greater neutralising antibody breadth will therefore be key to developing recommendations around the optimal development and deployment of such vaccines in the future.

Several studies have compared clinical protection from COVID-19 following different booster regimes. A major challenge in these studies has often been the use of different booster regimens during specific calendar time intervals, rather than in a (randomised) head-to-head comparison of two regimes. However, studies suggest that BA.1 is around 10% more protective than an ancestral booster^42^at protecting against symptomatic COVID-19, and up to 50% more effective at preventing severe disease^43,44^. Effectiveness of the BA.4/5 booster against symptomatic disease has been shown to have similar benefit compared to an ancestral booster (additional protection of BA.5 vaccine compared to an Ancestral vaccine was estimated at 8%, [95% CI 0-16%])^45^. In a comparison between boosters containing the BA.1 and BA.5 immunogens, little difference was discernible^46^. Thus, although there are fewer comparisons available, the data on clinical protection support the idea that updated boosters may provide a small increase in clinical protection; however, better prospective data collection on the effectiveness of different booster regiments would add valuable information.

It is important to note that in the four scenarios we could study to date, this trend has been consistent. Across the comparisons analysed, the estimated benefit of the updated booster immunogen ranged from 1.1 to 1.7-fold (Table 1). However, in future, the benefit of an updated booster immunogen is likely to depend on the antigenic distance between the vaccine immunogen and the variant that eventually circulates. It is possible that to date, by chance, or due to the selection pressure on virus, an improvement has been observed when using an updated immunogen because the variants that have emerged are progressively more distant from the originating variants^47,48^. It is possible that exceptions or variations to this pattern will arise as the antigenic diversity of the virus continues to increase. Therefore, future studies should assess whether the relative benefit of updating a booster immunogen can be predicted based on the antigenic relationships between the vaccine immunogen and the subsequent circulating variants.

This analysis has a number of important caveats. Firstly, we have only analysed in vitro neutralisation titres to variant immunogens, and not directly analysed the clinical protection provided by different booster regimens. However, despite this, the estimated increased odds of protection based on this analysis are in broad agreement with the studies outlined above.

Secondly, there were a limited number of studies^2,3,5,6,14,17,19,20,23,24^ that provided a head-to-head comparison of in vitro neutralisation titres elicited by different booster immunogens (Table 1). In a subset analysis, using only data from cohorts which included pre- and post-boost neutralisation titres for a future variant after boosting with *both* an old and updated immunogen (i.e. paired comparison in the same study), we found very similar results (update advantage of 1.37-fold [95% CI 1.03-1.82] Supplementary Table 6) despite the fact that this is based on less than half the data that was used in the full model. Thirdly, multiple different assays were used across the different studies to measure immunogenicity. We have accounted for this within our model by including an effect for the study from which the data was derived (a proxy for the assay used and the conditions under which it was run). However, lack of a standardised in vitro neutralisation assay remains a significant impediment to the ongoing comparisons of immunity following vaccination and infection^49,50^.

In addition, the data we have analysed comes from studies in which different variants were assessed at varying time points during the COVID-19 pandemic and after subjects had been exposed to different variants through natural infection. We attempted to account for these differences in out model by incorporating parameters to account for inter-study variation, and prior exposure history, but we cannot rule out an additional unexpected impact that we did not fully capture.

Publication bias also presents a potential challenge to interpretation of the results of booster comparison studies. For example, 9 studies^5–9,20–22,28^ that we identified were sponsored by the companies producing the vaccines. We therefore performed an analysis on a subset of studies^2–4,10,11,14,17,19,24^ that reported both pre- and post-boost neutralisation titres and had no pharmaceutical company involvement and found a similar benefit of updating the booster immunogen to that described above (update advantage of 1.39-fold [95% CI 1.04-1.84], Supplementary Table 7). Additionally, in our multiple regression analysis when we included pharmaceutical involvement as a potential factor, we found this variable was not significant.

Both these sensitivity analyses indicate that pharmaceutical sponsorship of studies is unlikely to have influenced our results. However, future studies should aim to provide unbiased analyses of neutralisation titres, preferably using standardised neutralisation assays, and directly comparing alternative booster regimes^51,52^.

Another major limitation of any retrospective study of COVID-19 variants and immunogenicity is the relatively short history of available data and the inherently unpredictable nature of future antigenic variation. Therefore, the predictions about the relative performance of an updated, compared to an older, immunogen can only be made based on the fairly limited number of prior immunogens that have been tested. The current “omicron” period of SARS-CoV-2 evolution seems characterised by the turnover of mutants displaying antigenic variation but with similar severity, however alterations to disease severity may emerge^53,54^. In addition, different variants have been co-circulating^55^, suggesting that no variant has a sufficient selection advantage to displace all the others.

Finally, there is no guarantee that a more antigenically escaped, more virulent, or more transmissible variant, will not arise in the future and alter the relative advantage of immunogen updates that we have identified. However, the findings here offer the best predictive tool to date that can be validated against future waves of variants and vaccine updates. Decisions on booster regimes will continue to be made based upon the currently available evidence. In this context we have shown that updating of the booster immunogen has, to date, increased neutralisation titres to subsequent circulating variants, and is predicted to have provided a modest increase in protection from COVID-19.

## Data Availability

Data and code will be made available following publication.

## Ethics statement

This work was approved under the UNSW Sydney Human Research Ethics Committee (approval HC200242).

## Authorship Statement

DC, MPD and DSK contributed to study design. DC, and AH performed the data extraction and curation. DC, AR and TES performed the data analysis. DC, MPD, DSK, SJK, AKW and JAJ contributed to shaping the direction of the work. All authors contributed to the writing and reviewed and approved the final report.

## Data and Code Availability Statement

Data and code will be made available following publication.

## Supplementary Materials

### Mixed Effects Model

As outlined in the main manuscript, the aim of our analysis was to consider pairs of potential immunogens, one designated as ‘old’ and one as ‘updated’, and compare their immunogenicity against a future variant, as outlined in in Figure 2 of the main manuscript.

We constructed a model that modelled either the fold rise in neutralising antibody titres or the absolute value of the neutralising antibody titres after boosting. The model included fixed effects for the booster immunogen (old or updated) and incorporated a random effect on the intercept with a grouping structure of the study from which the data came. The contribution of each data point was weighted by the square root of the number of subjects in the cohort from which it was derived, thereby giving more weight to larger cohorts.

The general form of the model of the antibody titres for cohort *i*, *T*_*i*_ is given by:

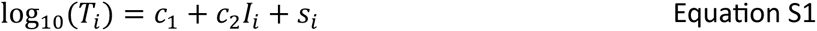

where

- *c*_1_is a constant representing the fixed effect for the intercept
- *c*_2_ is a constant representing the fixed effect for the difference between old and updated immunogens in the log10 of the fold rise in neutralising antibody titre
- *I*_*i*_ is a dummy variable determining whether the booster immunogen given to cohort *i* was old (*I*_*i*_ = old) or updated (*I*_*i*_ = updated).
- *s*_*i*_ is a random effect with a grouping structure of the study from which the data came.

We also constructed an extended model that accounted for the different exposure histories of subjects within the different cohorts. In this extended model, cohorts were paired if they were from the same study and had the same exposure history, but were given different booster immunogens. In addition to the effects described in the model in Equation S1, this extended model included a fixed effect for the exposure history of a cohort (average number of prior exposures within a cohort), a random effect on the intercept with a grouping structure on the pairing of the cohorts within a study (so that cohorts from the same study with the same exposure history, but with different booster immunogens, could be paired together within a study) and random effects on both the intercept and the contribution of the booster immunogen with grouping structures on the comparison that was being made (i.e. comparison number 1, 2, 3 or 4 from Figure 2).

The general form of the extended model for the antibody titres after boosting for cohort *i*, *T*_*i*_ is given by:

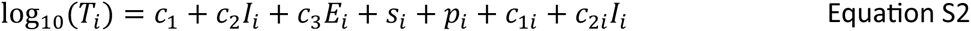

where

- *c*_1_ is a constant representing the fixed effect for the intercept
- *c*_2_ is a constant representing the fixed effect for the difference between old and updated immunogens in the log10 of the fold rise in neutralising antibody titre
- *c*_3_ is a constant representing the fixed effect for each additional exposure to the virus
- *I*_*i*_ is a dummy variable determining whether the booster immunogen given to cohort *i* was old (*I*_*i*_ = old) or updated (*I*_*i*_ = updated).
- *E*_*i*_ is a continuous variable representing the average number of prior exposures for the subjects within cohort *i*.
- *s*_*i*_ is a study-specific random effect.
- *p*_*i*_ is a pairing-specific random effect.
- *c*_1*i*_ and *c*_2*i*_ are comparison-specific random effects.

We defined the number of exposures for cohort *i*, *E*_*i*_, as the average of the number of prior vaccine doses received within the cohort (e.g. individuals receiving 2, 3 or 4 vaccine doses) added to their prior infection status. Prior infection status was defined as 0 (uninfected) or 1 (infected). In cohorts with mixed infection status, the prior infection status of the cohort was defined to be the proportion of the cohort that had undergone prior infection. If such information was not reported, the prior infection status of the cohort was set as 0.5.

### Statistical and model fitting

All analyses were conducted in R (v4.3.0) using library glmmTMB (v1.1.7). All reported p-values are based on the significance of the estimated parameters (Wald test), and the 95% CI are calculated directly from the estimated standard error of the parameters.

### Impact of boosting after multiple exposures to the virus – empirical data

A major challenge in comparing different booster regimens is that they were tested at different stages of the pandemic. For example, testing of the early VoC boosters generally occurred in subjects that had received only a primary vaccination course or had received a single booster dose. By contrast, comparison of the BA.1 and BA.5 bivalent booster vaccines occurred in a population that had up to 4 prior exposures (infections or vaccinations).

To determine the impact of prior exposures we initially grouped the extracted data by the number of prior exposures a cohort has previously experienced (prior to their boost). Where a cohort contained a mix of infected and uninfected subjects, and so had a non-integer mean number of exposures, we rounded the number of exposures to the nearest integer for this grouped analysis.

We observed a significant decrease in the magnitude of booster effect (for all variants) with increasing number of prior exposures (Supplementary Figure 2). We note that only that the *benefit* of a vaccine boost is reduced for subsequent exposures, not the absolute neutralisation titres. For each variant, considering the boosting of cohorts that have had primary vaccination and up to two additional exposures, an immunogen exposure resulted in higher post boost neutralisation titres (Supplementary Figure 3).

Despite this, when we consider the dataset depicted in Supplementary Figure 3 we observe that the geometric mean neutralisation titres against the XBB variant after 5 exposures (two-course primary vaccination and three additional exposures) are actually lower than the titres against BA. 1 after 3 exposures (two-course primary vaccination and one boost) (191 vs 604) suggesting that protective titres against the circulating variant are remaining relatively constant over time.

### Impact of boosting after multiple exposures to the virus – results of modelling

We next determined the contribution of exposure history to the rise in neutralisation titres following boosting from our mixed effects model. In this case we treated the number of prior exposures as a continuous variable. After accounting for different vaccine immunogens and comparisons, the number of prior exposures to a SARS-CoV-2 antigen still had a significant impact on the rise in neutralisation titres after boosting (p<.001). For each subsequent exposure to a SARS-CoV-2 immunogen, (up to the third exposure either from vaccination or infection) the magnitude of boosting decreased, on average, by 2.5-fold [95% CI 2.1-3.1] (negative slope of fitted model in Supplementary Figure 1B).

We observe that if this decrease were to continue, this would imply a fold rise of less than one, when boosting cohorts who had received four or more additional exposures after their primary series vaccination (i.e. a negative effect of boosting). Obviously, this is not biologically reasonable, and therefore this model is only applicable for the range of exposures over which we have available data with which to parameterise it. It is highly likely that, rather than having a fixed decrease of 2.5-fold in the magnitude of boosting for each exposure, this decrease may be smaller for each subsequent exposure. However, determining this will only be possible as more data becomes available for analysis.

## Supplementary Figures

**Supplementary Figure 1.**
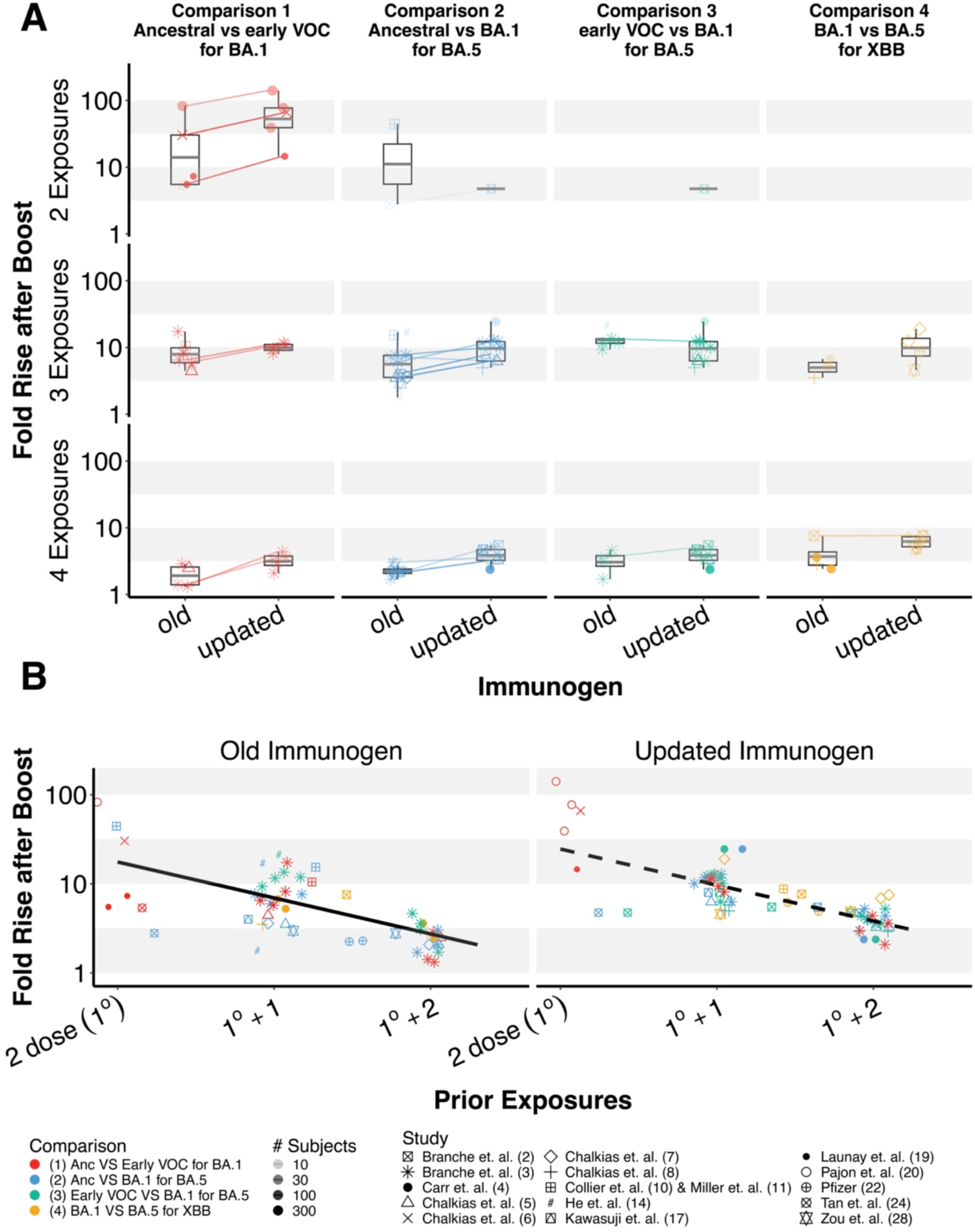
A) Representation of the data used to inform the mixed effects model of the fold rise in neutralisation titres after boosting with different immunogens. Data is stratified by the number of prior exposures (rows) and the comparison number (columns). Lines connect paired cohorts from the same study. (B) Model fits (lines) to data (symbols) showing the fold rise in neutralisation titres after boosting with an old (left) or updated (right) immunogen. Colours of symbols correspond to the different comparisons outlined in Figure 2.

**Supplementary Figure 2.**
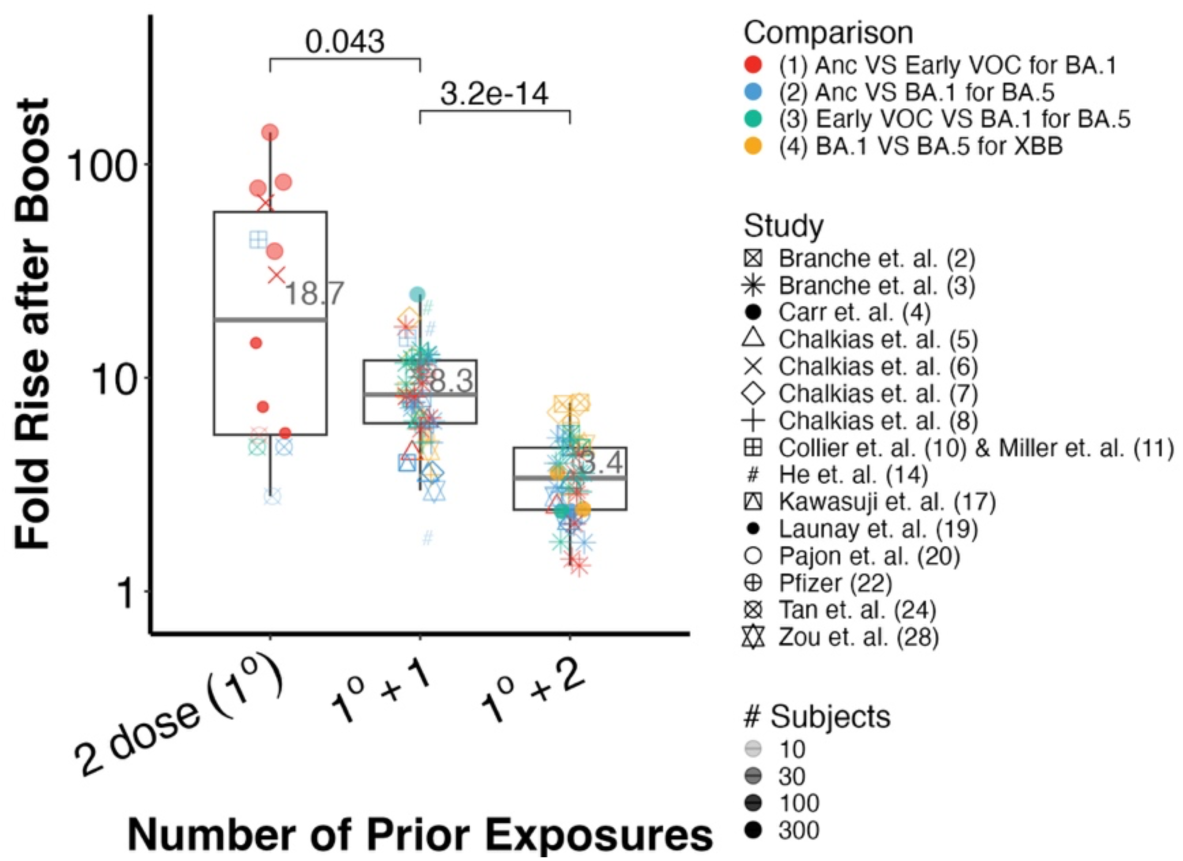
Fold rises in neutralisation titres after boosting with either an old or updated immunogen, stratified by the number of prior exposures within each cohort. Data are coloured by the comparison number to which they relate, and colours correspond to the comparisons depicted in Figure 2. Grey lines in the middle of the box plots (and numbers) indicate mean values (not median values). Comparison values across the top are p-values from unpaired t-tests.

**Supplementary Figure 3.**
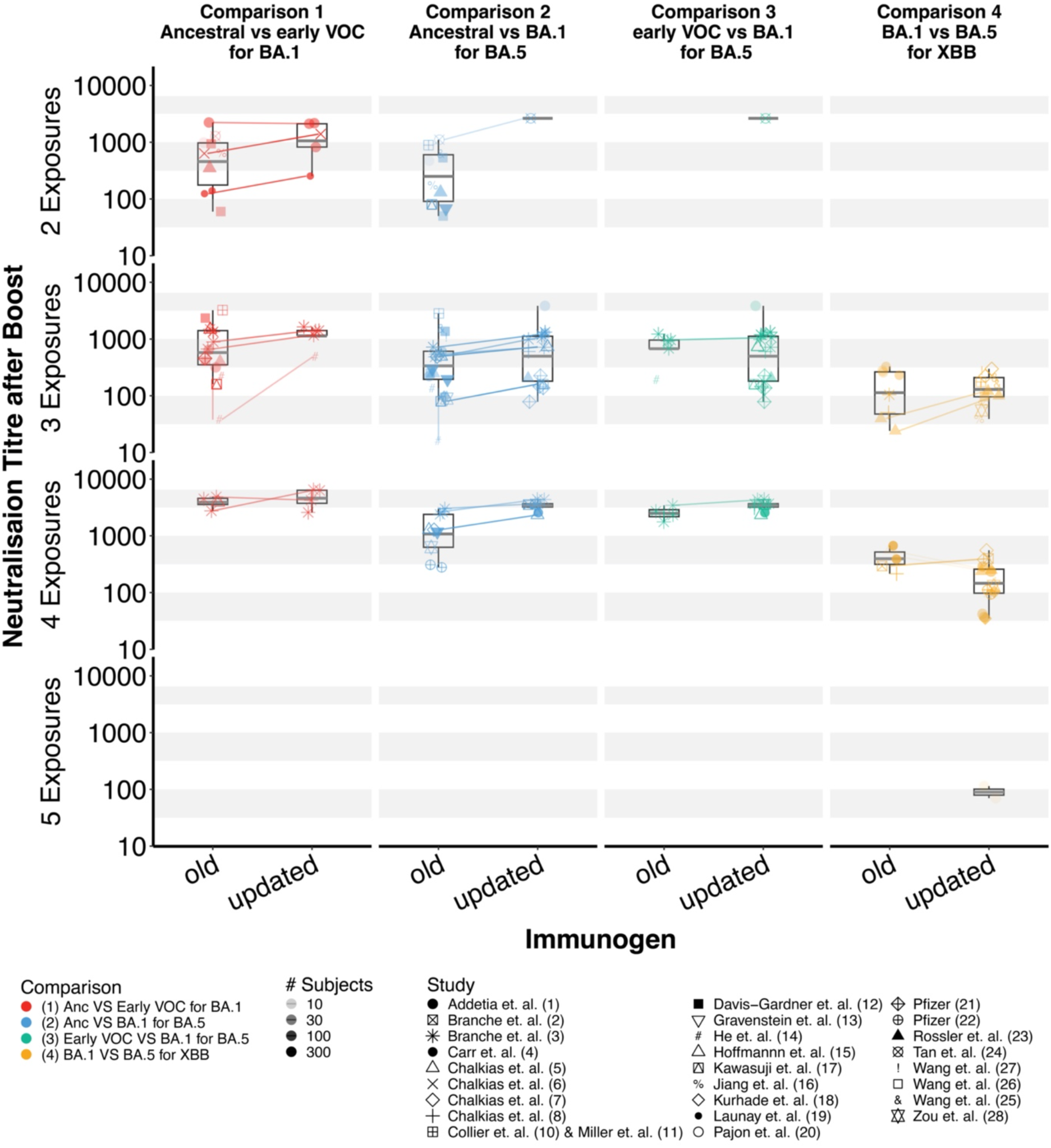
Representation of the data used to inform the mixed effects model of the absolute neutralisation titres after boosting with either an old or an updated immunogen. Data is stratified by the number of prior exposures (rows) and the comparison number (columns). Colours of symbols correspond to the different comparisons outlined in Figure 2. Lines connect paired cohorts from the same study.

## Supplementary Tables

**Supplementary Table 1:** Studies from which data were extracted to use in this analysis.

| Reference | Conflict | Has pre-boost data? | Immunogen | Relevant Variants | Number of Prior Doses | Prior Infectious Status | Relevant Data |
| --- | --- | --- | --- | --- | --- | --- | --- |
| <sup>1</sup> | None, however some authors are employees of Vir Biotechnology. | No | Ancestral, BA.1 bivalent, BA.5 bivalent | Ancestral, BA.1, BA.5, XBB | Multiple cohorts with different numbers of prior doses.<br>(i) Wu4 - 3 doses<br>(ii) Wu/BA.5 biv - mean 3.5 doses (range 2-4)<br>(iii) Pre-Omicron -Wu/BA.5 biv - mean 3.6 doses (range 3-4)<br>(iv) Omicron BT -Wu/BA.5 biv - mean 3.9 doses (range 3-5)<br>(v) Wu3 mono - 2 doses<br>(vi) Pre-Omicron-Wu3 mono - 2 doses<br>(vii) Wu/BA.1 biv - 3 doses | Multiple cohorts with different prior infection statuses.<br>(i) Wu4 - Uninfected<br>(ii) Wu/BA.5 biv - Uninfected<br>(iii) Pre-Omicron-Wu/BA.5 biv - Infected<br>(iv) Omicron BT-Wu/BA.5 biv - Infected<br>(v) Wu3 mono - Uninfected<br>(vi) Pre-Omicron-Wu3 mono - Infected<br>(vii) Wu/BA.1 biv - Uninfected | Figure 4 (A-B), cohorts (i-vii)<br>Data not extracted from cohort (viii) Omicron BT-Wu/BA.1 biv group as it was not clear whether the most recent exposure was an infection or a vaccination. |
| <sup>2</sup> | None | Yes | BA.1 bivalent, BA.5 bivalent | Ancestral, BA.1, BA.5, XBB | 3 doses | Mixed<br>*Monogram group* - Pfizer BA.1 bivalent group - 79% infected, 21% uninfected. Pfizer BA.4/5 bivalent groups - 75% infected, 25% uninfected.<br>*Duke group* - Mix of infected and uninfected, exact proportion not reported. | Figure 1(A-D), Table S6 and Table S7<br>Data from uninfected cohorts not used as no similar infected cohorts presented. |
| <sup>3</sup> | None | Yes | Ancestral, early VOC, BA.1 monovalent, BA.1 bivalent | Ancestral, BA.1, BA.5, XBB | 3 doses | Uninfected, Infected | Tables S6-S11 |
| <sup>4</sup> | None | Yes | BA.1 bivalent | Ancestral, BA.1, BA.5, XBB | 3 doses | Uninfected, Infected | Figure<br>Only data from uninfected and infected cohorts. |
| <sup>5</sup> | Supported by Moderna | Yes | Ancestral, BA.1 bivalent | Ancestral, BA.1, BA.5 | 3 doses | Uninfected, Infected | Figure 3, Figure S3, Figure S4<br>Only data from uninfected and infected cohorts |
| <sup>6</sup> | Supported by Moderna | Yes | Ancestral, early VOC | Ancestral, BA.1 | 2 doses | Uninfected<br>(up to 4% of a cohort has experienced prior infection, but these were still classified as uninfected) | Table S3 |
| <sup>7</sup> | Supported by Moderna | Yes | Ancestral, BA.5 bivalent | Ancestral, BA.5, XBB | 3 doses | Uninfected, Infected | Figure 3, Table S11<br>Only data from uninfected and infected cohorts. |
| <sup>8</sup> | Supported by Moderna | Yes | BA.1 bivalent, BA.5 bivalent | BA.5, XBB | 3 doses | Uninfected, Infected | Figure S5, Table S10<br>Only BA.1 variant data was used, as other data from this preprint is included in published form in reference <sup>7</sup>. |
| <sup>9</sup> | Supported by Moderna | Yes | XBB.1.5 monovalent, bivalent vaccine containing BA.5 and XBB.1.5 | Ancestral, BA.5, XBB | 4 doses | Uninfected, Infected | Figure 1 and Figure 2<br>Only data from uninfected and infected cohorts. Data only included in Figure 3 and not in the main analysis as no titres were provided after immunising with one of the relevant immunogens. |
| <sup>10,11</sup> | None | Yes | Ancestral, BA.5 bivalent | Ancestral, BA.1, BA.5, XBB | *Ancestral booster, uninfected cohort* - 2 doses<br>*Ancestral booster, mixed prior infection cohort* - mean 2.9 doses (range 2-3).<br>*BA.5 bivalent booster, mixed prior infection cohort* - mean 3.1 doses (range 2-4). | Uninfected<sup>11</sup><br>Mixed (33% infected, 67% uninfected) <sup>10,11</sup>. | Figure 1<sup>10</sup> and Figure S1<sup>10</sup><br>Figure 1(B-D)<sup>11</sup><br>(excluding WT and BA.5 data from Figure 1C and D) |
| <sup>12</sup> | None | No | Ancestral, BA.5 bivalent | Ancestral, BA.1, BA.5, XBB | 2 doses, 3 doses | Uninfected and Mixed For mixed cohorts:<br>*Ancestral booster cohort* - 27% infected, 73% uninfected.<br>*BA.5 booster cohort* - 17% infected, 83% uninfected. | Figure 1 (A - C) |
| <sup>13</sup> | None | No | Ancestral, BA.5 bivalent | Ancestral, BA.5 | 2 doses, 3 doses, 4 doses | Uninfected, Infected | Table 2 |
| <sup>14</sup> | None | Yes | Ancestral, early VOC, BA.5 bivalent | Ancestral, BA.1, BA.5 | 3 doses | Uninfected | Figure 2, Figure 3 |
| <sup>15</sup> | None | No | Ancestral, BA.5 bivalent | Ancestral, BA.1, BA.5 | 2 doses, 3 doses | Uninfected, Infected | Figure A, Figure B |
| <sup>16</sup> | None | No | Ancestral, BA.5 bivalent | Ancestral, BA.1, BA.5, XBB | 2 doses, 3 doses | Uninfected | Figure 3 |
| <sup>17</sup> | None | Yes, however pooled across all cohorts | Ancestral, BA.1 bivalent | Ancestral, BA.1, BA.5 | 2 doses, 3 doses | Uninfected | Figure 3B, Figure S2 |
| <sup>18</sup> | No, however one author received compensation from Pfizer for COVID-19 vaccine development. | No | Ancestral, BA.5 bivalent | Ancestral, BA.5, XBB | Multiple cohorts:<br>*Ancestral booster* - 3 doses<br>*BA.5 booster, uninfected* - mean 3.6 doses (range 2-4).<br>*BA.5 booster, infected* - mean 3.1 doses (range 2-4). | Uninfected, Infected | Figure 1 |
| <sup>19</sup> | No, however two authors are consultants to multiple vaccine companies and third is a consultant to Pfizer only. | Yes | Ancestral, early VOC | Ancestral, BA.1 | 2 doses | Uninfected | Figure 1 |
| <sup>20</sup> | Authors work for Moderna | Yes | Ancestral, early VOC | Ancestral, BA.1 | 2 doses | Uninfected | Figure S6 |
| <sup>21</sup> | Pfizer presentation | Yes | Ancestral, BA.1 monovalent, BA.1 bivalent | Ancestral, BA.1, BA.5 | 2 doses, 3 doses | Uninfected | Slides labelled CC8-CC19 |
| <sup>22</sup> | Pfizer presentation | Yes | Ancestral, BA.5 bivalent | BA.5, XBB | 3 doses | Mixed | Slide 9 |
| <sup>23</sup> | None. One author holds stocks in Pfizer. | No | Ancestral, BA.1 bivalent, BA.5 bivalent | Ancestral, BA.1, BA.5, XBB | 2 doses, 3 doses | Uninfected, infected | Figure 1, Figure 3 |
| <sup>24</sup> | None | Yes | Ancestral, BA.1 bivalent | Ancestral, BA.1, BA.5 | 2 doses, 1 dose | Mixed<br>*Cohort receiving Pfizer ancestral booster* - 11% infected, 89% uninfected.<br>*Cohort receiving Moderna BA.1 bivalent booster* - 32% infected, 68% uninfected. | Figure 3<br>Data from cohorts with only one prior vaccination was not included. |
| <sup>25</sup> | No, however one author is on the scientific advisory board of Janssen Biotech. and another has relationships with a number of medical development companies. | No | Ancestral, BA.5 bivalent | Ancestral, BA.5, XBB | 2 doses, 3 doses | Uninfected | Figure 2 |
| <sup>26</sup> | No, however one author is on the scientific advisory board of Janssen Biotech. and another has relationships with a number of medical development companies. | No | Ancestral, BA.5 bivalent | Ancestral, BA.1, BA.5 | 2 doses, 3 doses | Uninfected | Figure 1 |
| <sup>27</sup> | None | No | Ancestral, BA.5 bivalent | Ancestral, BA.5, XBB | 3 doses, 4 doses | Uninfected | Figure 1 |
| <sup>28</sup> | Supported by Pfizer | Yes | Ancestral, BA.5 bivalent | Ancestral, BA.5, XBB | 3 doses | Uninfected, Infected | Figure 1, Figure S1, Table S4<br>Only data from uninfected and infected cohorts. |

**Supplementary Table 2.** Parameter values for the mixed effects model described in Equation S2, that predicts the fold-rise in neutralisation titres following boosting and is fit only to the data shown in Supplementary Figure 1A from cohorts without prior infection.

| Dependant Variable | Parameter | Value | Std err | P-value |
| --- | --- | --- | --- | --- |
| Neutralisation titres after boosting | Intercept | 1.428 | 0.151 | <0.001 |
|  | Immunogen (updated vs older) | 0.187 | 0.072 | 0.009 |
|  | Exposure number | -0.654 | 0.169 | <0.001 |

**Supplementary Table 3.**
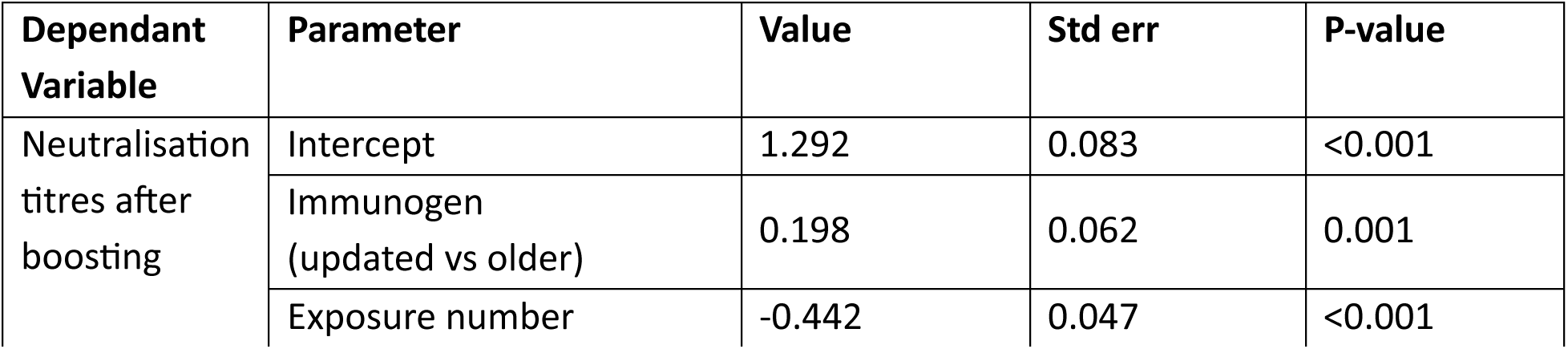
Parameter values for the mixed effects model described in Equation S2, that predicts the fold-rise in neutralisation titres following boosting and is fit only to the data shown in Supplementary Figure 1A from cohorts with a homogeneous prior infection status.

**Supplementary Table 4.** Parameter values for the mixed effects model described in Equation S2, that predicts the fold-rise in neutralisation titres following boosting and is fit only to the data shown in that did not include a comparison with a vaccine that contained an immunogen from an early VoC, i.e. based only on data from comparisons 2 and 4 from Supplementary Figure 1A

| Dependant Variable | Parameter | Value | Std err | P-value |
| --- | --- | --- | --- | --- |
| Neutralisation titres after boosting | Intercept | 1.099 | 0.115 | <0.001 |
|  | Immunogen (updated vs older) | 0.149 | 0.058 | 0.01 |
|  | Exposure number | -0.331 | 0.056 | <0.001 |

**Supplementary Table 5.** Parameter values for the mixed effects model described in Equation S2, that predicts the absolute neutralisation titres following boosting and is fit to the data shown in Supplementary Figure 3.

| Dependant Variable | Parameter | Value | Std err | P-value |
| --- | --- | --- | --- | --- |
|  | Intercept | 2.208 | 0.231 | <0.001 |
| Neutralisation titres after boosting | Immunogen (updated vs older) | 0.181 | 0.034 | <0.001 |
|  | Exposure number | 0.327 | 0.045 | <0.001 |

**Supplementary Table 6.** P Parameter values for the mixed effects model described in Equation S2, that predicts the fold-rise in neutralisation titres following boosting and is fit only to the paired data shown in Supplementary Figure 1A.

| Dependant Variable | Parameter | Value | Std err | P-value |
| --- | --- | --- | --- | --- |
| Neutralisation titres after boosting | Intercept | 1.278 | 0.176 | <0.001 |
|  | Immunogen (updated vs older) | 0.137 | 0.063 | 0.029 |
|  | Exposure number | -0.423 | 0.038 | <0.001 |

**Supplementary Table 7.** Parameter values for the mixed effects model described in Equation S2, that predicts the fold-rise in neutralisation titres following boosting and is fit only to the data shown in Supplementary Figure 1A that did not have pharmaceutical sponsorship.

| Dependant Variable | Parameter | Value | Std err | P-value |
| --- | --- | --- | --- | --- |
| Neutralisation titres after boosting | Intercept | 1.298 | 0.131 | <0.001 |
|  | Immunogen (updated vs older) | 0.142 | 0.063 | 0.025 |
|  | Exposure number | -0.504 | 0.055 | <0.001 |

